# Readability of selected governmental and popular health organization websites on Covid-19 public health information: A descriptive analysis

**DOI:** 10.1101/2020.06.27.20141770

**Authors:** Patricia Moyinoluwa Ojo, Tolulope Omowonuola Okeowo, Ann Mary Thampy, Zubair Kabir

**Affiliations:** School of Public Health, University College Cork, Cork, Ireland

## Abstract

**Background:** The U.S. Department of Health & Human Services (USDHHS) recommends that health material be written at or below a sixth-grade reading level to ensure readability. The aim of this study was to examine the readability of international and national health organizations on Covid-19 information in their websites employing a previously validated tool.

**Methods:** A purposive sample of publicly accessible governmental and popular international health organization websites was selected. The readability of the websites’ Covid-19 public health information was estimated using the previously validated SMOG readability formula, which determined reading level by correlating the number of polysyllabic words.

**Results:** Of the 10 websites included in the analysis, none had Covid-19 public health information at the USDHHS’s recommended reading level. The material ranged in reading level at undergraduate level or above.

**Discussion:** The findings indicate that the online Covid-19 materials need to be modified in order to reach recommended reading levels. This study can be of practical use to policy makers and public health government officials when designing, modifying, and evaluating Covid-19 materials. We recommend using simple, non-polysyllable words to ensure that Covid-19 public health information materials are written at the recommended reading levels.

## Introduction

Covid-19 is a new global pandemic affecting the respiratory system of an individual and ranges from mild to severe infections [1]. Vulnerable groups such as the elderly and individuals with medical illnesses (HIV, cancer and chronic respiratory disease) are prone to severity of the illness [1]. The virus outbreak began in Wuhan, China in December 2019, and the World Health Organization (WHO) declared the outbreak as a Public Health Emergency of International concern on 30^th^ January 2020 [2]. International health organizations, national governments and individual Departments of Health generally use internet as a reliable source to disseminate information to the public at a fast pace, thus creating public awareness of the disease and also profiling the risk characteristics of such health events. It has been documented that the sudden increase in Covid-19 cases in Wuhan City and in Hubei Province took a month to attract the United Nations’ attention on the overwhelming public health and health care services in China [3]. Importantly, the Covid-19 outbreak indicated that low health literacy among a population is an underestimated public health issue globally [4,5].

Effective communication along with daily updates through internet became the leading means of raising awareness around the world on Covid-19. Since these resources are directed towards every individual irrespective of their level of literacy, it is important that these resources are written in simple, plain language. Multilevel approaches like individual level communications, group level communications, community level social networking, policy level media activism, and population level media campaigns should be used to communicate at different levels of government [6]. According to The New York Times, the outbreak now has the highest prevalent cases globally which has caused unemployment rates to spike [7], in addition to other societal, health and psychological impact. Therefore, it is crucially important that unambiguous public health information be delivered using a reliable medium (websites) in an understandable context where misinterpretation is avoided on Covid-19 [8]. During this pandemic, research, effective communication, health literacy and health behavior have become crucial steps in attaining health and safety within the public [6]. As such, the U.S. Department of Health & Human Services (USDHHS) recommends that health material be written at or below a sixth-grade reading level to ensure readability [9].

Furthermore, it is important, collectively, that we create the degree of readability of Covid-19 online via governmental and international health organizations. Nonetheless, recent works on smoking cessation websites did not specifically concentrate on supporting evidence-based activities such as making health materials readable for those with poor literacy abilities [10-13]. Due to its new and developing features, minimal work was carried out. As such, this research builds on the findings of previous studies to assess the degree of online reading of Covid-19 materials from the websites of many governmental and international health organizations [10]. The readability of these resources can be measured using the Simple Measure of Gobbledygook (SMOG) which estimates the years of education an individual need to have to read a piece of text [14]. We set out to examine the readability of international and national health organizations on Covid-19 public health information in their websites employing this previously validated tool- the SMOG tool.

## Methods

### Readabilty Tool

There are several validated reading assessment tools such as the Simple Measure of Gobbledygook (SMOG) [14], the Flesch-Kincaid grade level, and the Gunning Fox index Because of a simpler interpretation and the easy availability of the SMOG tool, we prefered this to other readability tools. In addition, the SMOG tool was applied to an earlier study [10]. The SMOG readability is considered accurate and is highly correlated with other readability formulas; as such, SMOG has been recommended by the U.S. National Cancer Institute for determining the reading level of informational pamphlets about cancer [15]. Moreover, SMOG is one of the readability formulas recommended by the USDHHS and is commonly used by researchers and practitioners in medical and public health fields [16].

### Websites Selection

While Covid-19 has affected several countries, the country selection for this study was a purposive sample based on some pre-defined criteria: 1) population size; (2) Highest number of Covid-19 cases at the time of the study; (3) Internationally popular Covid-19 related health organiztions; (4) geographic representation; (5) China (the origin of Covid-19).

Therefore, the study included websites from federal health organizations, state health departments and local health offices with emphasis on Covid-19. Specifically, the Covid-19 websites of some individual state-health departments were included in the report, including the Health Service Executive (HSE) of Ireland.

We excluded websites if they (1) did not include Covid-19 information concerning the population are non-governmental organizations like private medical organizations and charity organizations (3) have weak webpages.

Based on the above criteria, the following websites were assessed for readability:

1. The World Health Organization (W.H.O)
2. The Health Service Executive (HSE), Ireland
3. The Centers for Disease Control and Prevention (CDC), United States
4. The European Centre for Disease Control and Prevention (ECDC)
5. The Government of Canada
6. The Federal Office of Public Health. Switzerland
7. The Public Health England (PHE), United Kingdom
8. The Ministry of Health, Italy
9. The Finnish Institute for Health and Welfare, Finland
10. The Department of Health, Australia.

## Analysis

The readability of the covid 19 information in each site was assessed using SMOG readability tool. SMOG test is a widely used readability test tool. It calculates the readability of the text using the number of polysyllabic words from the texts. Polysyllabic words are words that contain 3 or more syllables. A total of 30 sentences make the sample for calculating the readability. Three 10 sentence texts were taken from the beginning, middle and bottom portion of the websites and then fed into the online SMOG calculator. For texts with less than 30 sentences, the SMOG formula contains a factor to correct it. The texts were taken from the above 10 websites that provided health information on Coronavirus, symptoms, protection, prevention and treatment. SMOG measure indicates the minimum literacy level needed to understand the information provided in the websites. Following the measurment of the SMOG score for each website, the scores of each website were compared to the others to find their ease to understand by the public [17]

## Results

In this study, the selected 10 websites containing Covid-19 information was assessed for readability. The SMOG tool analysis (Table 2) determined the readability scores of these websites which ranged from a low score of 15.1 (CDC, Atlanta) to the highest score of 20.8 (Ministry of Health, Italy). Of the selected 10 websites in the analysis, none had Covid-19 health information at the USDHHS’s recommended reading level.

**Table 1:** SMOG readability score and the level of education needed ^[18]^.

| Level of education | SMOG score | Percentage of UK adults |
| --- | --- | --- |
| Entry 1 | 7-9 | 93% |
| Entry 2 |  |  |
| Entry 3 | 10-11 | 76% |
| Level 1 | 12-13 | 51% |
| Level 2 | 14-16 | 22% |
| Above level 2 | 17+ | <22% |

**Table 2:**
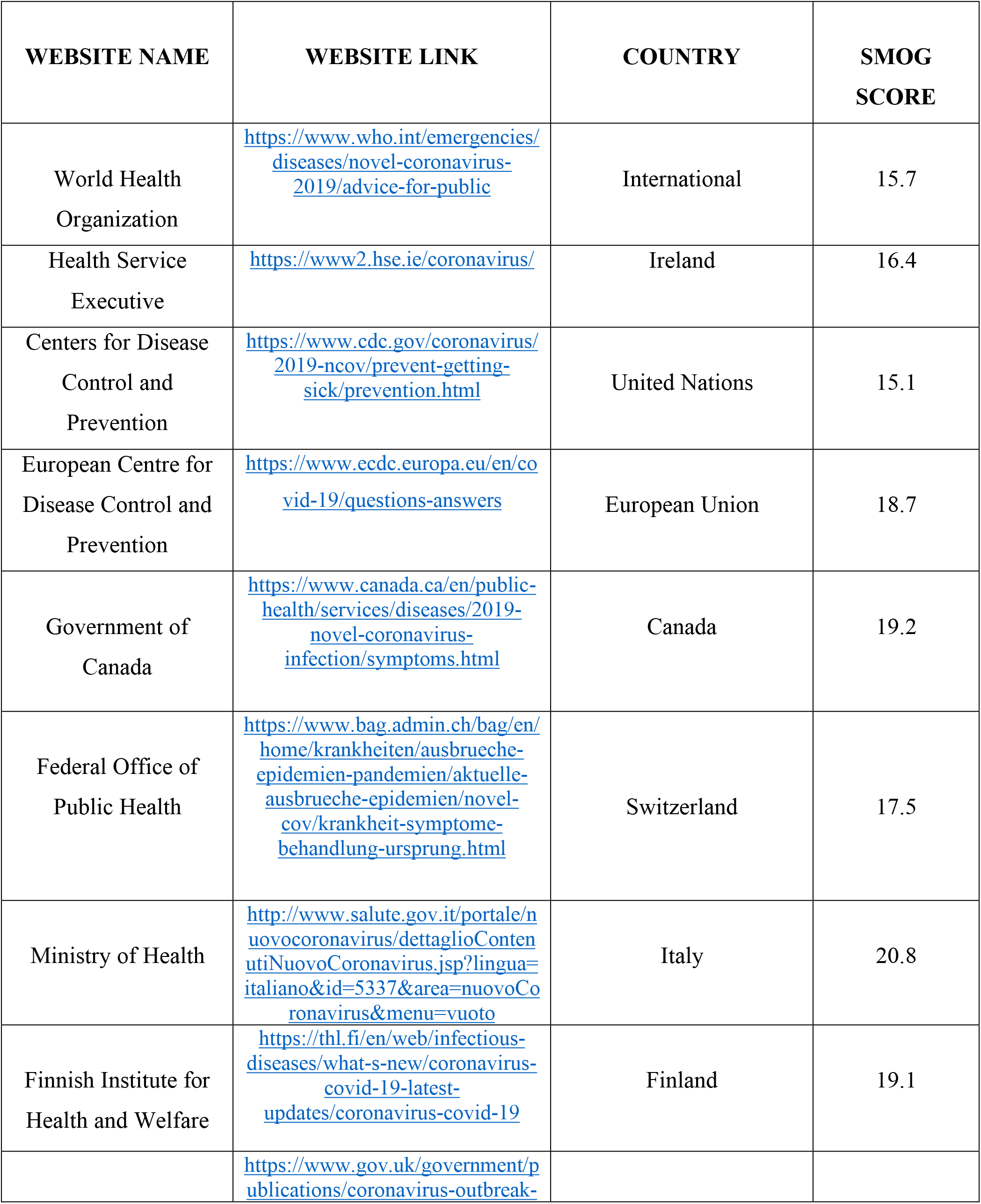
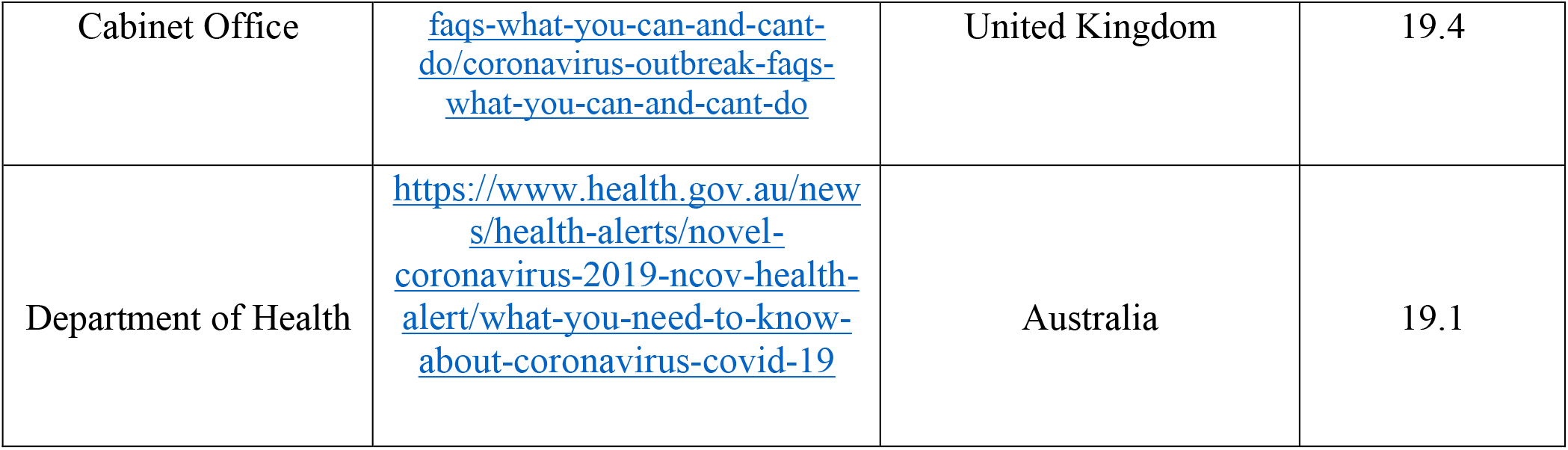
Reading level of COVID-19 information website.

The entire health information resource website analyzed was not written at an easy reading level - the average SMOG score was 16.2, and a SMOG score of 16.2 signifies a reading level for an undergraduate student. The National Health Service digital service manual in England recommend a SMOG reading score of 14 or less which indicates that the health information available is easy to read and understood by lay public, irrespective of their educational background.

## Discussion

This study evaluated the reading level of Covid-19 related public health information on selected governmental and popular health organization websites. Our SMOG assessment indicated a reading level which is equivalent to an undergraduate student level. In other words, none had Covid-19 health information at the USDHHS’s recommended reading level. This is an eye opener to public health officials to provide online education and health materials in compliance with the appropriate literacy levels for target population, which will help improve comprehension and better understanding for the target population or population at risk.

The Organization for Economic Co-operation and Development (OECD) Adult Skills Survey in 2012 reported that 1 in 6 Irish adults (17.9%) were at or below Level 1 on literacy scale. An individual at this level may be unable to understand written information. This survey assessed 6000 Irish people within the age range of 16-65 [19]. Governments have in effect encouraged their people to plan for a permanent, adaptable, and adaptive lifestyle due to the rapid changes in the world in terms of technology, political, economic, and social environment [19]. For instance, Ireland has introduced adult education through National Adult Literacy Agency (NALA) [8]. NALA is a charity organization that supports literacy through different online programs. Health literacy gives people confidence to make healthy, informed choices about health. The cheap procurement of electronic gadgets like mobile phones and computers along with easy availability of high-speed internet has helped people turn to online resources to collect information regarding all aspects of life. Online awareness and education have been more effective in case of emergencies especially in case on Covid-19 where any individual who owns an electronic gadget with internet can access all information that their respective governments have passed at their homes itself. This is particularly critical when there is a lot of ‘misinformation’ and a new wave of ‘infodemic’ [20].Writers can measure the reading level of information using computer generated reading tools which can be used to guide the reading level of health information [21].

## Methodological challenges

The information provided on the websites about Covid-19 was too long to understand. The use of conjunctions was used to connect sentences instead of constructing simple sentences. Most of the websites used medical jargons instead of plain and simple words which were not understandable for the public. Also, several websites were in their native languages thereby, excluding them from the study as they cannot be compared with other websites, for instance, in Chinese and in local Indian languages.

## Conclusion

The purpose of online resource website is to create an accessible and readable content for its target population. However, our study showed that this readability and comprehension are unintentionally neglected. In other words, none of the 10 websites analysed had Covid-19 public health information at the USDHHS’s recommended reading level. It is important that health literacy levels are accounted for while designing crucial public health messages on commonly used websites, which are largely accessible to lay public. Our study findings suggest that all Covid-19 and other health related websites should be written in simple/layman’s language, to substitute polysyllabic words with simple and plain words that are easier to understand and to process the public health information.

## Data Availability

Data will be available on request

## Ethical considerations

No personal data were identified or used, and therefore no instituional ethical approval was sought.

## Acknowledgement

This was part of a students’ group work project for the Master of Public Health (MPH) program in UCC, Ireland.

